# On the in vivo kinetics of gene delivery vectors

**DOI:** 10.1101/2022.02.11.22269834

**Authors:** Orestis Kontogiannis, Vangelis Karalis

**Affiliations:** School of Medicine, National and Kapodistrian University of Athens; Department of Pharmacy, School of Health Sciences, National and Kapodistrian University of Athens

**Keywords:** Gene delivery vectors, in vivo kinetics, viral vectors, non-viral vectors, adenoviral vectors

## Abstract

Gene therapy is the most promising strategy for treating a number of diseases at their most fundamental, genetic level, and it has a wide range of promising clinical and emerging preclinical uses in both the clinic and the laboratory. Gene therapy systems are composed of three fundamental components, with the delivery platform being responsible for the protection and successful delivery of the incorporated therapeutic nucleic acid sequences. A successful delivery platform is critical in the achievement of a therapeutic outcome, and an effective delivery platform is essential in achieving this. A variety of different gene delivery platforms - vectors - are evaluated in this dissertation in terms of their nature, mechanism of action, potential applications, and safety. Of particular importance is the evaluation of their post-delivery pharmacokinetic and adverse drug-metabolite profiles. The different types of vectors, including viral, non-viral, and alternative vectors, are discussed separately in each chapter, while important issues related to the incorporation of these vectors into clinical practice are discussed as well, including the topics of vector development and manufacturing, as well as the current regulatory landscape and efforts to improve it, and finally their prospects for the immediate future.

**Context:** Gene delivery vectors consist of a broad spectrum of natural or synthetically produced vehicles that represent one out of the three essential aspects of each gene delivery system, without which the successful and effective (in terms of the clinical translation) delivery of therapeutic nucleic acids to a diseased mal- or sub-functioning cell would be impossible.

**Objective:** The presentation and evaluation of the in vivo pharmacokinetic behavior of different viral and non viral gene delivery vectors including a wide review of their mechanism of action, possible safety concerns as well as the promise each holds for future applications.

**Data Sources:** A systematic literature search was conducted using the electronic databases PubMed, Google Scholar and Science Direct while also utilizing the Preferred Reporting Items for Systematic Reviews and Meta-analyses (PRISMA) guidelines. The keywords included in the research effort were the following: viral vectors, adeno-associated viral vectors, non viral vectors, oncolytic vectors, novel gene delivery vectors, pharmacokinetics of viral vectors, retroviral vectors, recombinant adeno-associated viral vectors, toxicity of gene delivery vectors, Vitravene, Oncorene, Approved gene delivery vectors.

**Study Selection:** Case studies and review articles published in scientifically accepted, high impact factor journals focused on gene delivery vectors and published in English between the years 1999 to 2021 in order to include the most significant findings in terms of both well established data through the years as well as the most recent breakthroughs in terms of preclinical and clinical application.

**Data Extraction:** Essential information were retrieved regarding the various types, behavior, mechanisms of action, safety and in vivo pharmacokinetic behavior of the most prominent viral and non viral gene delivery vectors.

**Results:** Of a total of 186 records a total of 36 full text articles were reviewed covering a total of 92 case studies and review articles on the topic of the pharmacokinetic behavior of gene delivery vectors including promising future considerations for their clinical use.

**Limitations:** The outcome of this review article was limited by findings that were shared between different articles published in a variety of literary platforms as well as from papers that lacked sufficient details in order to be included.

**Conclusion:** This scoping review has examined what is currently known and recently discovered regarding the wholesome of the aspects of utilizing specific gene delivery vehicles for a variety of different therapeutic purposes. Their nature, characteristics, as well as their individual action once inserted into the organism and/or in a variety of different in vivo experiments was examined and the implication of their use regarding their shortcomings and possible dangers, as well as their therapeutic advantages and probable future applications were weighted.

## 1 Introduction

The successful and targeted introduction and expression of nucleic acids, both DNA and RNA molecules, into a patient’s body cells to restore the function of a protein that is missing or defective due to a genetic disorder is referred to as gene therapy. The European Medicines Agency defines gene therapy as the use of biological medicinal products (such as nucleic acids, viruses, and genetically modified microorganisms) for therapeutic, prophylactic, or diagnostic purposes [1]. Due to limited experience with the technique and a lack of understanding of the mechanisms of some of the diseases mentioned above, gene therapy was previously only investigated for conditions where there was no other possible therapeutic approach to treat or alleviate symptoms [2, 3].

While using “naked” nucleic acids is thought to be the safest method for transfecting the desired cells, the interactions between the naked genetic material and cell surfaces, as well as the presence of several extracellular and intracellular barriers, make successful transfer unlikely [4]. In short, their anionic surface charge inhibits cell internalization due to repulsive forces (anionic cell surfaces) and facilitates their uptake by immune system cells, resulting in rapid clearance by the reticuloendothelial system (RES); intravenous administration renders them ineffective because they lack the stability to withstand rapid blood flow, and without an active targeting system, they are rendered ineffective [4, 5, 6].

Several solutions aimed at improving the delivery and targeting properties of naked nucleic acids have been investigated in the last decades in an effort to overcome these problems. These methods can be divided into two categories [4]: a) Physical and mechanical methods and b) Gene delivery carriers, also known as gene delivery systems or vectors, are used.

The first group of methods includes electroporation (the application of an electric field to increase the permeability of the cell membrane), sonoporation (the use of ultrasonic frequencies to alter the permeability of the cell membrane), optoporation or optical transfection (the use of light to achieve internalization), gene gun, and microinjection. The ultimate goal of these techniques is to deliver exogenous nucleic acids to the appropriate host cells. Although their use is preferable to the administration of naked genetic material under various conditions, the above methods are associated with high costs in the majority of cases and appear impractical in most gene delivery applications [4, 7-9].

In terms of gene delivery systems, this is still the most common method for successfully transfecting genetic material into mammalian cells (transfection efficiency is measured by the proportion of encapsulated nucleic acid that can correct the function of a diseased cell). These systems are the result of the combination of three fundamental components: a) a plasmid-based gene expression system – controlling the therapeutic gene’s function within the targeted cell, b) a therapeutic gene – one that is specific to the disease being treated and encodes the correct and functional protein, c) a gene delivery vector – is in charge of delivering the gene expression plasmid to the desired location (specific organ and cell type) within the human body [2,4,6].

The vectors, which are used to successfully deliver therapeutic nucleic acids into infected cells, are divided and classified into viral vectors, which include recombinant viruses and are also known as biological nanoparticles, and non-viral vectors which include synthetic vectors forming complexes with genetic material [10]. The aim of this study is to provide an overview on the kinetics of viral and non-viral gene delivery vectors and present some successful applications on the market.

## 2 Literature search

### 2.1 Methods

A systematic literature search was conducted using the electronic databases PubMed, Google Scholar and Science Direct while also utilizing the Preferred Reporting Items for Systematic Reviews and Meta-analyses (PRISMA) guidelines in order to complement the research effort. In order to gain a thorough overview of the research field at hand the research methodology was structured in two distinctive – consecutive parts with the first phase being the research for and selection of papers to include in the review (including the type of papers used, of which time period and which keywords to research for), while the second was the classification and taxonomy of the papers previously gathered. The initial research was conducted between January 2021 – July 2021, and updated October 2021 – November 2021. A thorough search was conducted in the published literature to extrapolate data for pharmacokinetic analysis of a variety of viral, nonviral, and alternative vectors for gene delivery. The goal was to gather as much data as possible to help form a holistic, spherical view regarding vectors for gene delivery, from their nature, mechanisms of action, safety concerns, development and scale-up to their regulatory landscape and, most importantly, their pharmacokinetic behavior.

Regarding the data extraction process, the extracted variables occurred by following a standardized data extraction template, extracting information on the identification data (first author, publication year and journal of publication) type of the study, the study characteristics and location, the publication year and the methods used.

### 2.2 Search strategy

A thorough search of the published literature was conducted to extrapolate data for pharmacokinetic analysis of a variety of viral, nonviral, and alternative vectors for gene delivery. The goal was to gather as much data as possible to provide a holistic, spherical view of vectors for gene delivery, from their nature, mechanisms of action, safety concerns, development and scale-up to their regulatory landscape and, most importantly, their pharmacokinetic behavior. The Preferred Reporting Items for Systematic Reviews and Meta-Analyses (PRISMA) statement [11] was followed when conducting this review (Fig. 1).

**Figure 1.**
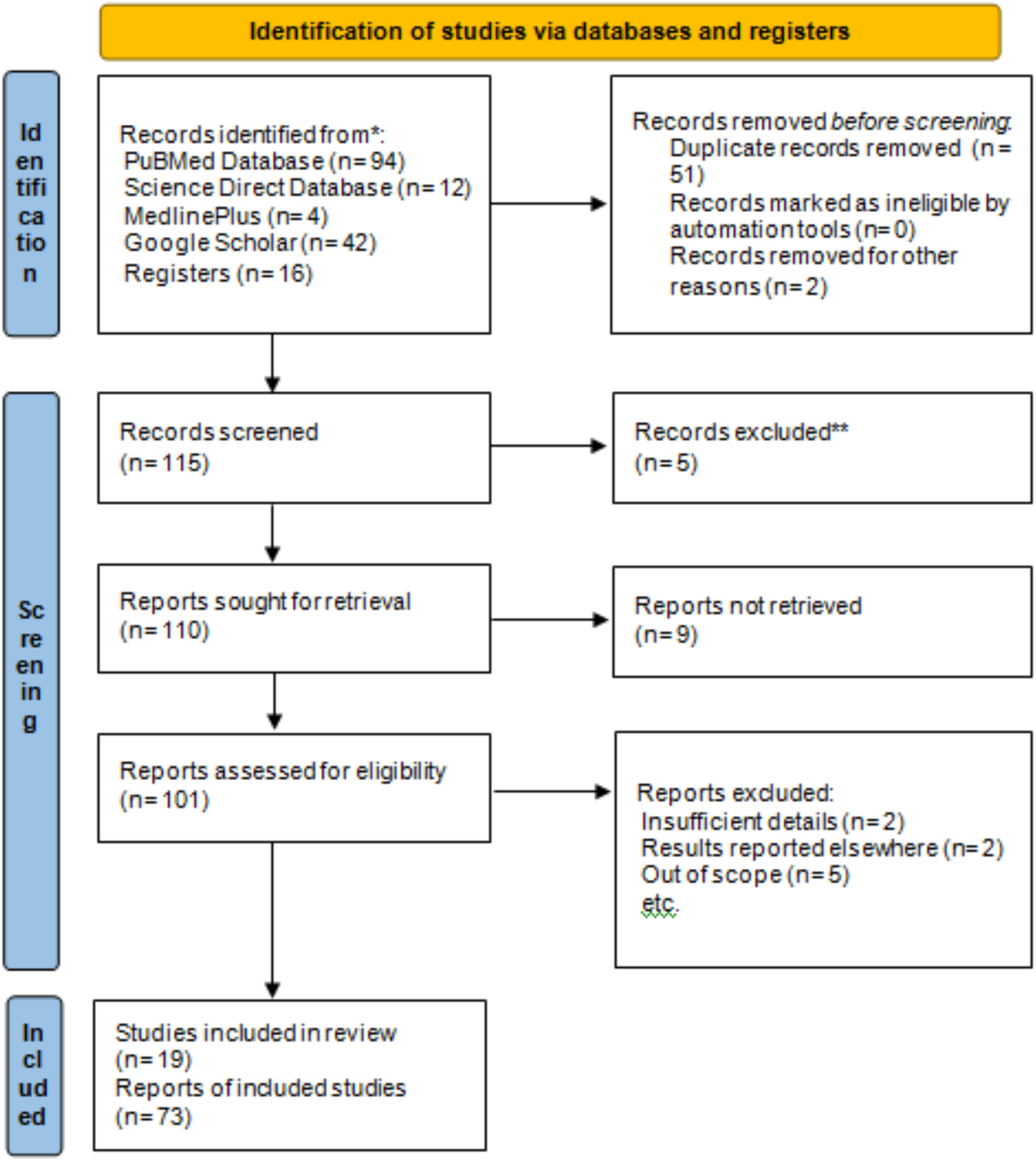
Study search criteria and selection process using the PRISMA method.

The following combination of terms was used to identify and collect the papers required for the first phase as previously mentioned: viral vectors, adeno-associated viral vectors, non viral vectors, oncolytic vectors, novel gene delivery vectors, pharmacokinetics AND viral vectors, non viral vectors AND pharmacokinetics, retroviral vectors, recombinant adeno-associated viral vectors, toxicity AND gene delivery vectors, Vitravene, Oncorene, Approved gene delivery vectors, gene delivery vectors AND FDA* OR EMA.

### 2.3 Selection Criteria

For the identification of relevant papers, the following criteria were applied.

#### Inclusion criteria

- Case studies, clinical trials and reviews
- The paper had to include numerical data regarding the nanoparticle physicochemical properties in relation with the pharmacokinetic variations they provoke.
- Data that can be used to create viable pharmacokinetic models
- Publishing data not prior to 1999
- Language: English

#### Exclusion criteria

- Newspaper articles
- Conferences abstracts and letters

## 3 Key findings

### 3.1 Viral vectors

#### 3.2.1 Types

Viral vectors are used to deliver therapeutic genetic material into target cells. These vectors deliberately lack some of the genes involved in the reproduction process that their wild-type precursor viruses possessed [12, 13]. The first viruses accessed were retroviruses, which have been used to develop several products with high therapeutic efficacy, and adenoviruses (Ad), which were first studied around the same time [14]. These vectors, as well as adeno-associated viruses, herpes simplex viruses, oncolytic viruses, human foamy viruses, vaccinia viruses, and lentiviruses (usually derived from HIV), monopolize the majority of scientific interest in viral vector studies (Fig. 3) and are found in the majority of clinical trials and publications [2, 14].

Despite their many benefits, the majority of viral-based therapies lack some of the characteristics that would allow gene therapy to achieve some of its ultimate goals, such as the possibility of a single dose life-long treatment and ease of incorporation into clinical practice.With the advancement of vector production, hybrid forms of the above vectors that combine several advantages of the different vector types are being synthesized.

The type of genetic material introduced into host cells and whether the therapy is mediated by DNA (deoxyribonucleus) or RNA (ribonucleus) molecules are two other broad categories of viral vectors. Oncolytic viral vectors, a third category, are primarily used to treat various types of cancer. Some RNA viruses use DNA intermediate molecules for replication and can thus be used to deliver double-stranded nucleic acids. DNA-based vectors persist in vivo for a longer period of time and are integrated directly into the cell’s genome. Viruses in this category include lentivirus, poxvirus, adenovirus, adeno-associated virus, retrovirus, human foamy virus (HFV), and herpes virus. RNA-based vectors are typically shorter-lived, with therapeutic effects lasting only a limited time. Viruses in this category include human foamy virus and oncoretrovirus [15].

In contrast to mammalian host cells, which receive protein synthesis information from double-stranded DNA molecules, retroviruses carry their genetic information in the form of RNA molecules. When viral nucleic acid enters cells, the expression of the newly added viral “genes” does not always begin immediately. Internalization occurs via the recognition of specific receptors found on the surface of specific cell types, implying that their infectivity is site-specific and thus more limited [15]. To address this issue, a technique in which the virus’s endogenous envelope proteins are replaced by proteins derived from other viruses can be used, resulting in a wider range of infection. These are known as pseudotyped viruses (a notable retroviral example is the VSV Gpseudotyped virus). Depending on the circumstances, this method can also be used to achieve the opposite result, i.e., to limit the range of infection to specific cell types [10, 15].

Recombinant adeno-associated viral vectors (rAAV) began to displace conventional adenoviral vectors over time and are now widely regarded as the most advantageous delivery platforms, providing a better clinical outcome in many gene therapy applications [16, 17]. The external morphology and crystallinity of these engineered delivery systems are similar to that of adeno-associated viruses (same capsid), but they lack segments of the natural viral genome. Their main advantages are in the internalization process and the extent to which the transported nucleic acids are expressed: [16-18]

#### 3.2.2 In vivo kinetics

According to preclinical data, cell internalization of adenoviruses, and thus most adenoviral vectors, occurs in vitro via endocytosis, specifically via the mechanisms of pinocytosis and clathrin-dependent endocytosis. Most of the time, and primarily because of the nanometer size range of these viruses, this mechanism is preferred over others that are more prevalent with larger molecules, such as phagocytosis. This internalization pathway involves uptake after interaction with the host cell’s clathrin coating, followed by transport through the intracellular matrix via clathrin related vehicles (CCV) to the lysosomes [13, 15]. Internalization through nucleus pores is mediated by adenoviral hexon proteins and dynein light intermediate chains, as well as their interactions with microtubules, followed by the inevitable “hijacking” of cellular reproductive function [15, 19, 27]. A vector can be synthetically modified to change its cellular preference, allowing for targeted delivery directly to the desired tissue while also facilitating viral transport through more inaccessible anatomical regions of the human body such as the brain, central nervous system (CNS), lymphatic system, and synovial fluid [13, 20].

After internalization, adenoviruses remain in a dormant state for a variable amount of time, causing no harm to the host cell[21]. During the latent period, the virus is stable, despite the fact that each serotype can achieve a plethora of integrated and episomal forms that attach to different regions of different chromosomes.

Adenoviral vectors are known to rapidly induce immune responses that activate antigen-presenting cells such as macrophages and dendritic cells, which process and then present the internalized antigen-in this case, adenovirus-for recognition when administered systemically(intravenous injection) [13, 15]. Depending on the dose administered and the type of tissue, this results in biphasic toxicity, both acute with rapid onset of symptoms and late effect toxicity. Some of these viruses lack the ability to accumulate and efficiently penetrate the desired cell membranes, and thus require the assistance of another virus, known as the helper virus, to facilitate internalization. The herpes simplex virus, for example, is used as a helper virus for the dependoparvovirus [15, 18]. The use of recombinant vectors (Fig. 2) is made possible by the use of packaging cells, as the virus’s genomic sequence has been altered by deleting certain pathogenicity-associated regions and then integrating the therapeutic nucleic acid. These packaging cells can “package” the vector containing the desired gene into a transport vehicle [22]. Once packaged and formulated, these vectors exploit the virus’s inherent ability to infect host cells and, once genetically integrated, exploit the cell’s mechanisms for their own replication. Finding the right balance between the molecular design of the recombinant vector cell line and the final genetic sequence, the type of promoters used, and the optimal regulatory mechanism for the expression of all of the above factors is one of the most important aspects of hoping for a successful therapeutic effect [18, 22]. When compared to other delivery systems, recombinant vectors have been shown to achieve higher infection rates [22].

**Figure2.**
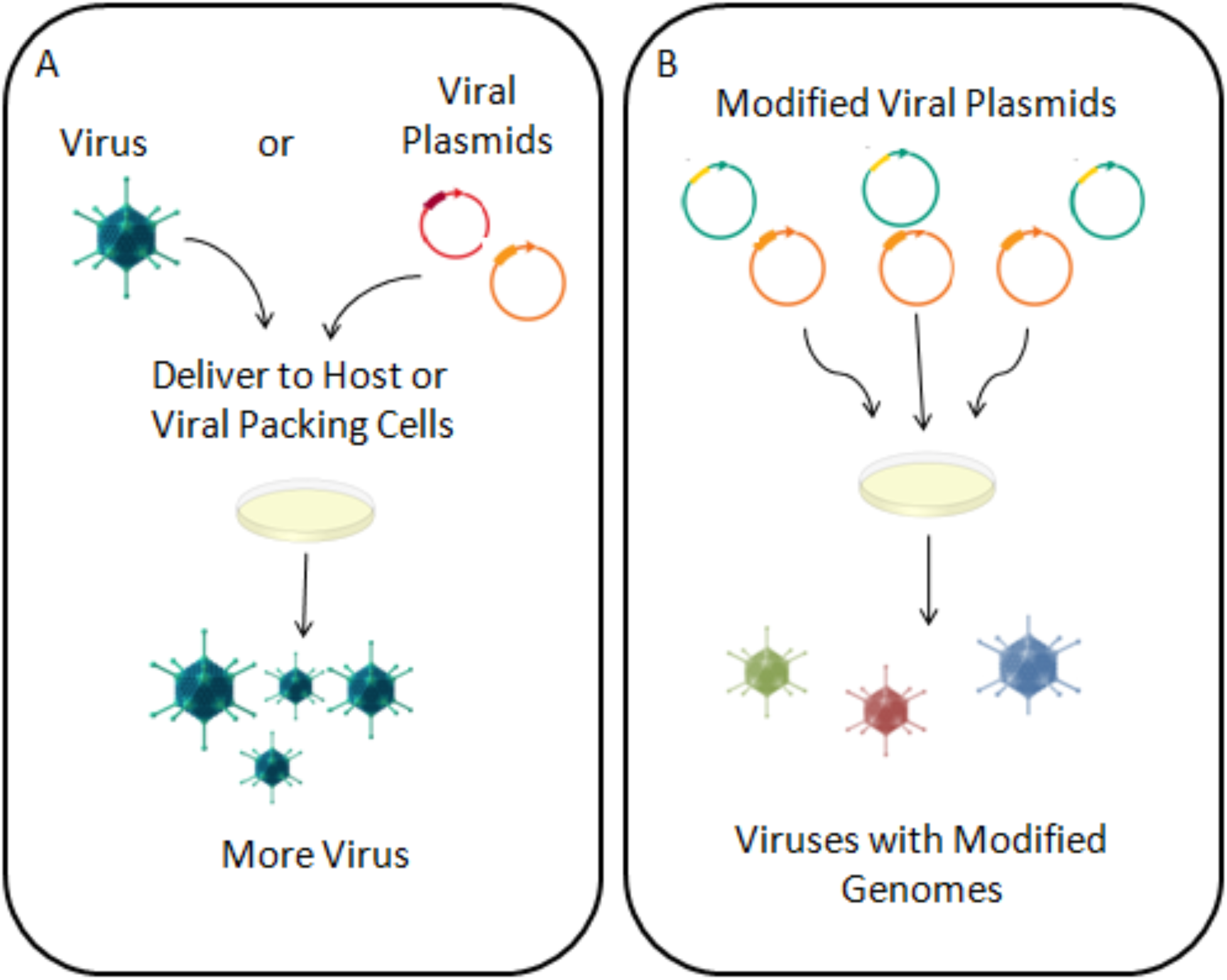
Delivery mechanism of viral vectors.

**Figure3.**
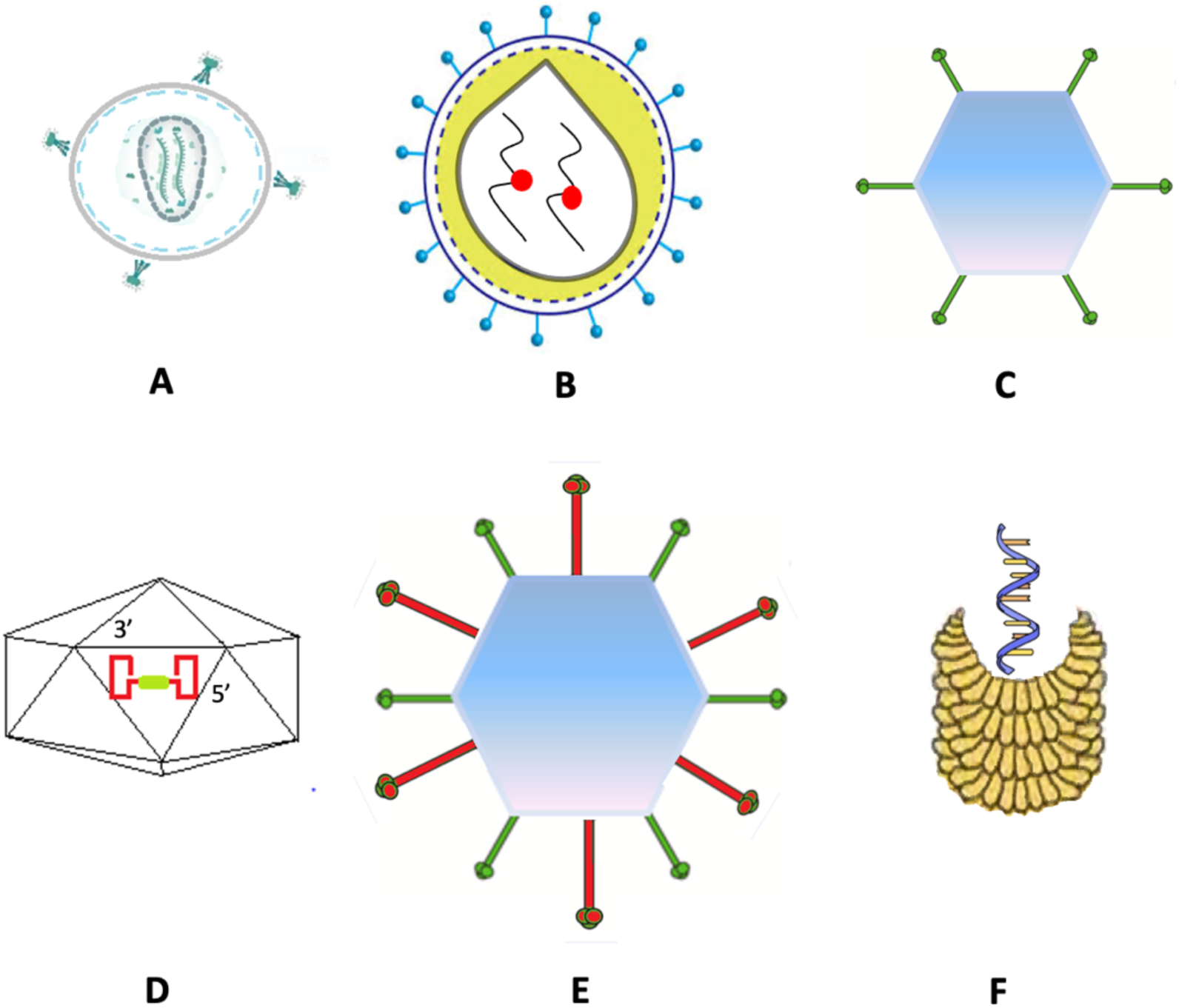
Viral vector types. A:Retroviruses; B: Lentiviruses;C: Adenoviruses;D: Adeno-associated viruses; E: Hybrids;F: Plant viruses.

As part of their life cycle, retroviruses (including lentivirus and foamy virus) can integrate their genetic material directly into the DNA of the infected cell. Retroviridae viruses have the enzymes required to fuse the incorporated nucleic acid with the genome of the host organism. Because each cell division contains the genetic material of the provirus integrated, ready to be replicated, transcribed, and translated alongside the DNA of the original cell, this mechanism has the potential to achieve lifelong expression of the therapeutic nucleic acid. This genomic integration has the potential to influence several aspects of retrovirus behavior, including their overall availability to cells of interest, the rate of unwanted accumulation in nontargeted tissues, and distal toxicity [13, 23]. In terms of the overall immunogenic profile of the system, as well as the stability and rate of degradation of ribonucleic acid in the final formulation, RNA-based delivery systems outperform. In summary, over the last few decades, a number of promising platforms for RNA delivery in mammalian cells have been developed, each with its own mechanism of action, including: [2] i) Naked syn-mRNA (usually administered via intravenous injection or in a few preclinical studies in aerosol format against lung disorders – mostly useful in vaccine development), ii) Viral or non-viral RNA encased within viral particles, and iii) Self-amplifying RNA replicons – relatively short RNA molecules synthesized in the laboratory.

#### 3.2.3 Pharmacokinetic aspects

Viral vectors have pharmacokinetic properties that differ markedly from those of conventional small molecules (molecular weights up to several hundred daltons). Therefore, information from other indications of the viral vector, such as the precise formulation and purification processes, titration methods for viral normalization, incubation time, performance characteristics, and inherent batch-to-batch variability, is critical and can be used to guide pharmacokinetic analysis.

With respect to the resorption pattern of most viral vectors, the route of administration has a significant impact on the internalization process by altering the environment to which the system is exposed, the intracellular and extracellular barriers that the vectors must overcome, and the primary tissue and cell type to which they are exposed. In general, topical delivery has been shown in several experiments to limit the overall availability of the system, as well as its ability to penetrate the various epidermal barriers and successfully reach the inner layers of the living epidermis and eventually the dermis to reach the blood capillaries. These findings have been confirmed in both animal models and human skin substitutes (3-D cell layers) as well as xenografts (immune-compressed mice that serve as receptors for human genetic material to develop human tissues including cancerous tumors) and show limited promise for applications in diseases such as autoimmune diseases like psoriasis and melanoma [13, 24].

Several inhaled gene therapies for transporting oligogucleotide sequences have been used to treat various respiratory diseases, with promising results in terms of low extrapulmonary organ transfection and increased absorption in the lung (in the majority of airway tissue cells, from the upper airways and bronchioles to deeper lung tissue such as the alveoli - although smaller size is required for such deep penetration) [13, 24]. The first-pass effect, which occurs primarily in the liver but also in the epithelial cells and epithelial cell membranes of the gastrointestinal tract, is one of the reasons why oral delivery platforms for gene therapy, including viral vectors, are not among the most intensively studied areas. Preclinical studies have been conducted in mice as well as larger animals such as dogs (although lumen permeability is usually overestimated) and pigs, but to date no significant study has progressed beyond phase I. In preclinical studies, in vivo experiments are performed in animal species to assess the process of delivery of the administered dosage form into the systemic circulation, and in most cases they are performed to collect important toxicological data. Animal models such as rodents (murine, fecine, cricetine, and musteline) and various primate families can simulate human lumen function and provide useful information on the fraction of the initial dose absorbed; however, no model has shown data correlating with human gastrointestinal activity in terms of drug bioavailability after the first-pass effect. When the delivery system is administered intravenously, intradermally, or subcutaneously, human body activity is best mimicked in animal models [13, 24].

The goal of biodistribution studies is to identify the primary and possible secondary organs in which a viral vector may distribute and accumulate after administration, as well as the cellular targets and types that may be found in these organs. These preclinical studies may be limited to various in vitro or in vivo animal models, or additional experiments may be performed, such as direct injection into organ targets, e.g., by parenchymal injections, with the notable example of injection into the spine using serotype 9 adenoviral vectors. Among the rAAV vectors, those of serotype 2 in particular have been extensively researched and studied (rAAV2). They are found in significant amounts in the liver, followed by distribution in other organs such as the spleen, smooth muscle, striated muscle and kidneys. Using rodent models, the researchers discovered that recombinant adeno-associated viral vectors of serotype 6 (rAAV6) accumulate primarily in the liver, but also in equal amounts in the spleen, although after extraction of the animal organs, a nonspecific distribution pattern was observed that followed traces of the vectors found in several other tissues. Since other similar types of the same family (e.g., serotype 1) are considered as alternative vectors that have not yet shown promising results in gene delivery, they have not been studied in detail with respect to pharmacokinetics [23, 25, 26].

The probability of distribution to sperm is an important issue when working with adenoviral vectors and recombinant adeno-associated vectors. In gene therapy applications, the intention is not to treat germline cells as purely somatic cells. In several preclinical studies using in vivo animal models (rodent, rabbit, dog), adenoviral DNA was detected in animal testes, both in the basement membrane and in the interstitial space. Encouragingly, no evidence of vector internalization was found in spermatozoa, suggesting that vectors may not be able to interact with the cellular receptors of this cell type [27].

When the excretory pathway of viral vectors was examined, traces of AAV2 were detected in the kidneys and thus in the bladder, suggesting that urinary excretion is a possibility that should be considered. Following inhalation and intranasal administration of the vector, excretion via the liver and gall bladder, as well as via feces (if saliva is ingested), is usually observed, while other routes, such as menstrual blood and sputum, are considered likely and are currently under investigation [26]. Decisions made during the formulation process that affect the distribution and accumulation profile of viral vectors can impact the route of excretion, as changing characteristics such as carrier size (Table 1) can lead to faster or slower excretion, as can excretion through different organs (kidney, lung, liver, and spleen have the ability to process substances of certain size ranges) [13]. Clamping the portal vein, hepatic artery, and bile duct allows vectors that are processed and excreted through the liver to bypass hepatic clearance and circulate longer in the bloodstream [28]. Attachment of polyethylene glycol to viral capsid molecules is a technique that can be applied to any vector to successfully increase its blood circulation, meaning that regardless of the external morphology of the particles, an efficient degree of coverage has been created to avoid immune recognition and opsonization [13, 29]. Adenoviral vectors, modified vaccinia virus, and herpes simplex virus type 1 are examples of replication-deficient viral vectors that persist in vivo for a short time before being eliminated by the organism [23].

**Table 1.** Vector drainage route depending on size.

| Route of clearance | Viral Vector’s size |
| --- | --- |
| Renal Clearance (Kidneys) | <5nm |
| Hepatic Clearance (Liver) | >100nm |
| Spleen (RES/MPS) | >20nm |
| Pulmonary Clearance (Lungs) | <20nm (Disse space) or <100nm (Lumen) |

Adenoviral vectors, on the other hand, are unable to maintain long-term stable circulation due to the nature of human physiology, resulting in virions with short half-lives. The reticuloendothelial system is responsible for the elimination of the vectors, leading to their uptake by Kupffer cells in the liver and eventual hepatic clearance [30]. Normal mice have a residence time in the blood compartment of about 2 minutes, and while low concentrations of particles (3×103) are rapidly eliminated, higher doses of viral particles can overcome this natural filtration (Kupffer cells) and show a non-linear dose-response relationship in hepatic gene transduction. Interestingly, the blood compartment had the lowest yield for recovery of adenoviral nucleic acid copies once inside the organism in several experiments [23].

Regardless of the type of viral vector, the origin of the progenitor virus should always be considered. If the vector is derived from a human virus, it is likely that most people will have come into contact with the progenitor virus at some point in their lives and will have developed antibodies to it. It is expected that the residence time and half-life in the body will be significantly reduced in this case [29].

### 3.2 Non-viral vectors

#### 3.2.1 Types

Non-viral gene delivery technology has emerged as a result of recent advances in vector synthesis, allowing the development of novel molecules and the application of techniques with the same degree of cellular transfection as viral vectors [2, 10]. This field of study and application encompasses a wide range of synthetically produced molecules, each with its own advantages and disadvantages in terms of efficacy, loading capacity, safety, immunogenicity and tolerability, ease of manufacture and scaling, and overall production cost [13]. Non-viral vector technology is generally based on the use of liposomes or polymers, and these two types of molecules are the primary non-viral vectors used in the majority of current applications. Liposome-based vectors allow gene transfer through direct interaction with nucleic acid sequences, resulting in the formation of transfer complexes known as lipoplexes, which are formed electrostatically. Due to the anionic charge of DNA, cationic liposomal molecules can spontaneously and rapidly bind to it upon contact. Polymeric vectors work on the same principle, but they form electrostatic bonds between the polymer chains and the DNA, which are called polyplexes. The positively charged polymer molecules have the added advantage of being able to hide the negative charge of the transferred nucleic acid, which facilitates recognition by the immune system, and of being able to “compress” larger DNA sequences into smaller, more compact structures (thus improving the internalization profile).

To date, most cationic polymeric non-viral vectors are polyamines such as poly(L-lysine), known as PLL, poly(ethylenimines), poly(amidoamines), poly[2-(dimethyamino)ethylmethacrylates] and chitosans, each of which has its own positively charged structures and binding moieties [2, 4]. The ease and simplicity of the gene transfer process, while maintaining generally high efficacy, is a major advantage of using the above molecules. The chemical (positively charged regions, length of linker sequences) and structural-geometrical properties of the polymers used determine the effectiveness and transfection ability of the complexes formed [2, 22, 31]. Since most of these vectors have been synthetically modified and engineered, they exhibit significant differences in terms of preparation, properties, and efficacy in various applications, making comparison with viral vectors difficult.

DOTMA and its ester analogs DOTAP and SAINT -2 are the most commonly used lipidic vectors. They all have amphipathic structures (i.e., they contain both polar and nonpolar parts in their structure, resulting in different solvent affinity) and have capabilities and properties similar to those of naturally occurring phospholipids [32]. Polymers are classified as either naturally occurring or synthetically produced, with both types being used in a variety of biomedical applications. Animal collagen, chitin, chitosan, keratin, silk, and elastin or plant starch, cellulose, and pectin are compounds with high biocompatibility and biodegradability, but lack some key properties of synthetically produced ones, such as increased thermal stability and “smart” mechanical properties. Synthetic polymers, on the other hand, can usually be fine-tuned and manufactured in a variety of shapes and sizes with desirable morphological properties to meet the requirements of the therapy in question, and offer advantages in stability and transfection that natural polymers, which are more difficult to manufacture, do not.

Other molecules (Fig. 5), such as dendrimers, polyphosphoesters, and peptides, interact with nucleic acids via their amines or ammonium ions due to their electrostatic nature and are among the most promising candidates for nonviral gene delivery from a variety of cationic polymers, with many studies in recent years focusing on the biocompatible polymer PLGA or poly(lactic-co-glycolic acid) [13, 22].

As for the third category of nucleic acid complexes (nanocomplexes), nanoparticles are a promising material to successfully deliver a variety of nucleic acids intracellularly. These vectors can easily overcome the cellular barriers of plasma membranes of eukaryotic cells with sizes ranging from 10 to 1000 nanometers (we accept structures with diameters larger than 100 nm, but with properties related to their small size or/and with one or more external dimensions or a nanoscale internal structure) [13, 16].

Gene delivery vectors based on cyclodextrins have also been studied and tested in vivo. These natural cyclic oligosaccharides are highly biocompatible and can form bonds with anionic nucleic acids to form novel complexes (CDplexes). The US FDA generally considers these molecules to be safe. In fact, they are classified into three groups based on the bonds they form: α-, β-, or γ-cyclodextrins (alpha, beta, gamma). They can be cationically charged by substituting small positively charged compounds, and have been shown in several applications to increase the mucosal permeability of a variety of encapsulated compounds [22].

#### 3.2.2 In vivo kinetics

Various physical, artificially induced methods can be used to internalize and permeate therapeutic nucleic acids across the plasma membranes of eukaryotic human cells for non-viral mediated gene delivery. Physical methods include needle injection, ballistic DNA injection, sono-poration, photo-poration, magneto-infection and hydro-poration. Chemical methods using compounds that can form sterically stable bonds with DNA or RNA molecules and facilitate their targeted transfer and internalization are also widely recognized as successful delivery methods. In addition, engineered vectors are thoroughly studied and developed. They are based on the formation of electrostatic forces that generate nucleic acid complexes capable of circulating in the bloodstream for extended periods of time before being internalized by the endocytosis mechanism. Their “intelligence” in terms of microenvironment identification is based on a variety of techniques, including the incorporation of functional groups and surface ligands, as well as the fact that the nature of these electrostatic bonds exhibits media-dependent sensitivity and therefore behaves differently in different parts of the human organism where different conditions prevail (e.g., pH differences). Scientists are particularly interested in built-in targeting moieties at the outer periphery of the vector that are covalently-non-ionically-coordinated with DNA complexes and provide general facilitation in terms of attachment to non-viral delivery platforms without compromising their original properties [2, 4].

The overall morphology and colloidal stability of the system can be modified to our advantage in various applications where lipid-nucleic acid complexes are formulated to mediate gene transfer by incorporating neutrally charged helper lipids such as DOPE and DOPC, whose easily modifiable and sterically stable geometries allow them to assume different shapes. Thus, these vectors condense the encapsulated genetic material to a greater extent, resulting in greater overall uptake. Calculating the packing parameter of the system (P = V/aLc, where: ratio of the volume of the hydrophobic tail v, product of the area of the hydrophilic head group a, length of the hydrophobic tail Lc), for a value of P of 1, we conclude that the area occupied by the hydrophobic part of the molecule is greater than the area occupied by the water-soluble head group, which means that the formulation is hydrophobic. Using the same methodology as before, other values of P are indicators of different morphologies, revealing the different physical and thermodynamic properties of the product produced [33].

Since most of the above delivery platforms form electrostatic complexes with the genetic material, the ability to form stably bound, irreversible aggregates with a variety of plasma proteins, as well as the formation of so-called “hard” coronae, especially with the initially more abundant plasma proteins, is an important consideration in terms of their interactions and mechanism of action after administration. The use of stealth technology and molecules that control their positive charge (polyanions such as polyacrylic acid, polyvinyl sulfate, and polyethylene glycol) leads to systems with enhanced gene expression and preferential long-term, gradual production (possibly due to increased circulation times) of therapeutic proteins and thus improved clinical outcomes, where u [13, 27, 34].

#### 3.2.3 Pharmacokinetic aspects

Intravenous injection, topical application, mucosal administration, *per os* administration and finally in aerosol form (inhalable) via pulmonary administration are the primary methods of delivering such a system into the host. The first-pass effect, which occurs primarily in the liver but also in the epithelial cells and epithelial cell membranes of the intestinal mucosa and prevents the dissolved portion of the drug from being fully bioavailable, is the reason why oral administration is avoided in gene therapy applications. As a result, a significant portion of the therapy is lost after administration, and therapeutic efficacy suffers significantly. After insertion, the vector must overcome the initial obstacles to its efficacy, which include enzymatic degradation, blood flow and pressure, complement-mediated recognition and clearance, and opsonization. Research shows that neutral particles tend to aggregate in physiological salt concentrations, leading to reduced efficacy, cytotoxicity problems, clotting problems, and pulmonary embolism. Cationic particles are more stable in solution, but once mixed with human plasma, they tend to bind with abundant blood proteins such as albumin and are then easily recognized and removed from circulation, while aggregation phenomena can be observed in a time-dependent manner depending on the vector and proteins of the complex [22, 33]. The vector used in targeted cancer therapies should be designed to overcome additional barriers present in the tumor extracellular matrix, such as the hypoxic and pH-depleted environment, as well as the abnormally shaped microvasculature surrounding cancer cells, which has poor transport and lymphatic drainage properties and a high degree of leakage (6,5-6,8). These changes can be taken into account in the design of the drug delivery platform (morphology, chemical properties, targeting ligands, binding stability), but if not, they lead to ineffective and unstable drug delivery[13, 22]. When products designed for cancer treatment with non-viral vectors are administered systemically, they show beneficial properties in terms of disease treatment, since by achieving stable circulation within the blood for long periods of time and a high permeability profile, the vectors can gain access to both the original tumor site and any other secondary metastasis by reaching essentially intact any vasculitis [22].

The ability to infect the lung via respiration is a major advantage of non-viral vector technology. Aerosol - spray products can effectively infect large numbers of lung cells at high concentrations with virtually no side effects, and they do not even require minimally invasive methods such as bronchoscopic administration [22]. Due to their small size, non-viral vectors are internalized and penetrate the cell membrane via the endocytosis (Fig. 4) mechanism, where they are transported through lysosomes and primary and late endosomes, being exposed to a variety of pH environments ranging from extracellular pH values of 7 to about 5 or even slightly lower. Endocytosis can occur via specific protein binding (caveolae or clathrin-dependent) or nonspecific internalization, the latter generally favoring a pathway that avoids lysosome transport. Non-viral vectors must overcome lysosomes, as well as endosomal escape, which is known to be a major limitation to successful nuclear internalization. Polymer-based vectors and polyplexes, when exposed to various intracellular enzymes, have been shown to be readily degradable and unstable in these low pH environments, resulting in their conversion to smaller polymers upon bond breakage and failure to protect their genetic cargo. To address this problem, positively charged polymers with high buffering capacity and structural flexibility can rapidly exit the endosome, possibly via the so-called “proton sponge effect” (flow of ions and water across the endosomal membrane). Another advantage of charged molecules is their ability to interact with plasma membranes without the need for functionalizing/targeting ligands [13, 35]. The formation of a lipid/graft that mediates the endocytosis process is required for the majority of polyplexes to be internalized. Positively charged lipids, such as some of these cationic polymers, have the advantageous ability to bind to proteoglycans (nuclear receptors of a variety of viral and pathogenic cell internalizations) and use this binding to mediate cell transfection [35]. After successful subcellular escape, the nuclear membrane and transport through nuclear pores become the next barrier to effective therapeutic gene expression. This step is thought to occur through the action of two separate mechanisms, the first being direct diffusion through the pores and the second being mitosis, which aids the passage of larger molecules through the pores. The efficiency of cellular transfection is to some extent dependent on the cell cycle stage of the infected cell, with the S and G2 phases being more beneficial in terms of gene expression than the G1 phase, suggesting that the presence of a mitotic environment is preferable (the closer we get to phase M, the more membrane degradation facilitates internalization) [12, 36, 37].

**Figure4.**
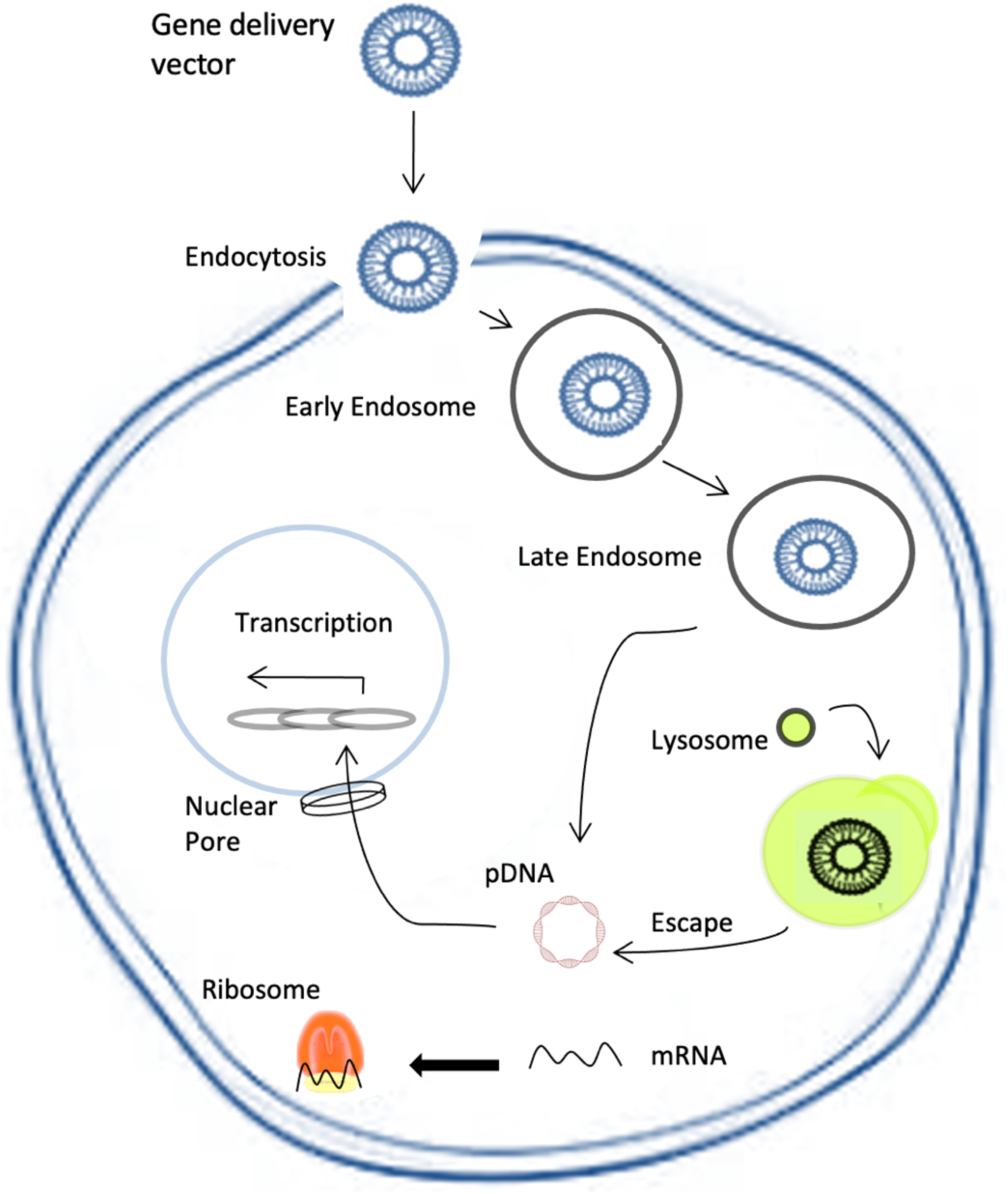
Delivery mechanism of non-viral vectors.

**Figure5.**
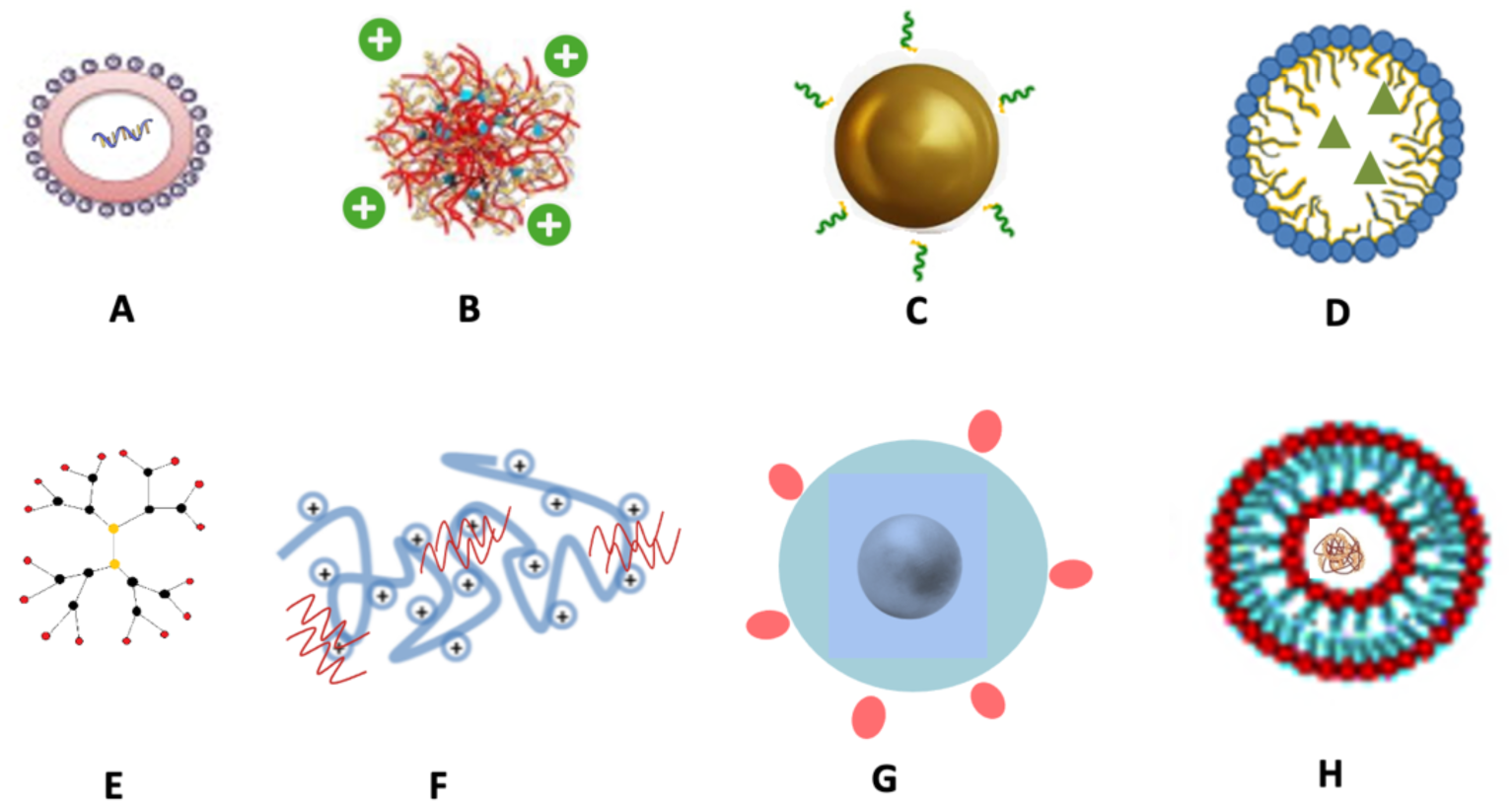
Non-viral vector types. A: Cationic Lipids; B: Cationic Polymers;C: Inorganic NPs;D: Lipid Nano Emulsions; E: Dendrimers;F: Chitosan;G: Peptide Based; H: Polyethylenimine.

Regardless of the site of initial injection, polymeric or lipidic - DNA complexes are rapidly transported through the venous network and taken up by the heart after systemic administration. Because the lungs are composed of the smallest capillaries in the body, only small-diameter molecules can pass through, while larger vectors are intercepted, which explains the increased risk of pulmonary embolism due to restricted blood flow. This phenomenon also explains why gene transfection in lung tissue leads to high expression levels, as a large number of particles accumulate there and are transported into these cells in large quantities after cardiac passage [22]. Delivery systems with low plasma stability and higher probability of degradation tend to release (“leak”) some of their genetic cargo before reaching the target site, which poses an interesting problem in terms of studying the pharmacokinetic behavior of potentially freely circulating nucleic acids. While the packaging and geometry of encapsulated DNA or RNA plays an important role in their ability to circulate in the bloodstream, as well as in their half-life and degradation profile, these molecules generally cannot remain intact and retain their properties in the host microenvironment unless they are protected [27, 37].

With respect to the elimination profiles of nonviral vectors (metabolism and excretion), several physicochemical properties determine their ability to be degraded and metabolized by different organs within the organism, as well as their ability to be filtered and excreted or to accumulate in different tissues after administration without being degraded. The overall hydrodynamic size of a vector is one of the most important factors affecting its ability to be transported to RES or other excretory organs for processing and excretion. Several studies have shown that the kidneys in particular are an important excretory pathway for non-viral gene therapy vectors via the renal clearance mechanism. In general, vectors with diameters up to 25 nanometers show a faster clearance profile with residence times of approximately 1.5 hours, whereas systems with diameters from 50 to 250 nm were eliminated more slowly [22, 37]. It is important to note that non-viral vectors are subject to the same size limitations as viral vectors when it comes to clearance through various organs of the human body [38].

As mentioned earlier, charged lipoplexes and polyplexes tend to interact with various blood plasma proteins, and this binding can lead to the formation of a protein corona, which in turn triggers rapid removal of the complex from the bloodstream via nonspecific interactions with components of the RES system. The mononuclear phagocyte system is essential for the elimination of recognized molecules by the Kupffer cells of the liver. Of course, as with other delivery platforms, PEGylation is sufficient to mask these molecules by hiding their charge and preventing complex formation by efficiently covering the reactive regions of the vector, resulting in increased circulation time, prevention of opsonization, and improved chance of reaching the cellular site of interest [38].

### 3.3 Applications

The list of products already approved by both the FDA and the EMA, the majority of which target somatic diseases, indicates that gene therapy research and applications continue to focus primarily on the treatment of somatic diseases. Three approaches are commonly used in the treatment of such diseases: in vivo delivery, ex vivo delivery and finally in situ delivery:

i. In vivo delivery, where- the therapeutic nucleic acid is delivered directly into the target tissue
ii. Ex vivo delivery, where the genetic material is implanted into the bone marrow of the target tissue, cultured and manipulated in vitro, avoiding any immunological reaction (the majority of cells remain non-viable after implantation)
iii. In situ, where the genetic material is introduced directly into the target tissue (as the name suggests, in situ studies are confined to a specific site within the whole organism and are usually considered intermediate between in vivo and in vitro) [2, 39].

Formivirsen, marketed as Vitravene® [40], was the first gene therapy drug approved by the FDA in 1998, whereas in China, the first commercially available gene therapy product Gendicine® (recombinant adenoviral vector expressing p53, rAd-p53) was approved by the State Food and Drug Administration of China (SFDA) in 2003 and is now known as National Medical Pro [13]. Two years later, in 2005, the USFDA approved Oncorine®, the first oncolytic viral drug, for the treatment of nasopharyngeal carcinoma. In August 2018, Onopattro® (Patisiran), a drug whose mechanism is based on gene silencing, became the first RNAi product to be approved by both the EMA and FDA, while Kymriah® (tisagenlecleucel) and Yescarta® (AxicabtageneCiloleucel), were the first two CD19-targeted chimeric antigen receptor (CAR) [1].

The FDA has approved the products Kynamro® (mipomersen), Imlygic® (TalimogeneLaherparepvec), Luxturna® (VoretigeneNeparvovec-rzyl), Spinraza® (nusinersen), and Exondys 51® (eteplirsen) in the last decade, while Strimvelis® (autologous CD34+ cells transduced to express human aden [36] In addition to the FDA, EMA, and China, the Philippines Bureau of Food and Drugs (BFAD) approved Rexin-G® (Mx-dnG1), a gene therapy indicated for both soft tissue sarcomas and osteocarcinomas, in 2007 [1]. It employs a retrovector, and recent studies have shown that Rexin-G® monotherapy against metastatic tumors has promising safety and efficacy profiles. Neovasculgen®, a plasmid-based drug, was also approved by the Russian Ministry in 2011. Collategen® (BeperminogenePerplasmid) is Japan’s first gene therapy product (MHLW) approved for the treatment of critical limb ischemia (CLI). It is critical to note that conventional treatments are ineffective in more than 20-40% of CLI patients in the United States. Finally, Heath Canada approved Tegsedi® (inotersen), an antisense RNA drug, in October 2018 [1].

### 3.4 Safety

Jesse Gelsinger ‘s death in 1999 shocked the entire gene therapy community because he was the first person to die from complications related to clinical trials of a gene therapy product. His death was attributed to the lack of an adequate adverse effect reporting system, which resulted in two previous cases being dismissed, and to Mr. Gelsinger’s high ammonia levels, which should have excluded him from the trials. Due to safety concerns, the technique was discontinued in the early years of the twenty-first century, particularly in the United States and Europe [1, 41].

Due to the nature of the treatment, which uses recombinant protein infusions that must penetrate diseased cells, and some additional properties that the system is supposed to possess that can lead to severe toxicity effects if misjudged, gene therapy applications carry a certain percentage of risks. The technique may lead to unfavorable effects caused by the overall system, i.e., the synergistic combination of all its components, or by the individual interactions of the vector or therapeutic nucleic acid with the human organism. Consequently, for specifically designed non-viral vectors (typically at the nanometer scale), toxicity studies should examine the system as a whole, rather than relying on prior knowledge of the safety of individual components [3, 13].

If the ultimate goal is to produce enough of a therapeutic protein to circulate systemically in the body, the type of cells transfected may be unimportant as long as an efficient clinical effect is achieved. In other cases, the success of the technique depends on getting the drug to specific cells in the body and allowing it to attach to their surface so that it can penetrate the cell membrane before releasing the therapeutic genetic material in the cytoplasm, nucleus, or a specific subcellular organelle. In these two cases, either the nonspecific distribution of the drug or the incorrect application of the chosen active targeting technique could lead to unwanted toxicity and accumulation in healthy tissues. In addition, the responses to therapy, such as stimulation of the immune system, are not fully understood because the interactions of these nanoscale systems with antigen-presenting cells cannot be adequately monitored [13].

While each product has its own synthesis and different physicochemical (size, shape, surface charge), pharmacokinetic (ADME profile) and thus toxicological properties, a common risk of each system, whether with a viral or non-viral vector, is the potential development of drug resistance, especially in cases where multiple doses are required following long-term treatment modules, leading to toxicity[42, 43]. The two main pillars for approval and evaluating the success of clinical and preclinical studies in regulatory science for the development and use of any new drug are efficacy and safety and, of course, the balance between the two. This rule does not apply to gene therapy products. In the following section, we will consider vectors that have higher transfection efficiency than others and thus produce more of the desired therapeutic molecule, but are considered less suitable for various applications due to their slow immunogenicity [4].

## 4 Future perspectives

Currently, there are many gene therapy applications (Tables 2, 3) that are very promising and resourceful in treating various inherited or acquired genetic disorders, such as infectious diseases, cancer, neurological disorders, mutations associated with cardiovascular abnormalities, and the majority of other single-gene or more complex disorders [6, 44]. In addition, gene therapy is gradually transferring its tools and knowledge to other scientific fields, such as bionanotechnology, to study and understand the basic cellular mechanisms and to produce specific therapeutic proteins when needed. The overlap of these fields has led to gene delivery platforms created using nanomedicine (nanotechnology applied in medicine) that have shown promising results in their ability to treat lethal genetic disorders [6].

**Table 2.**
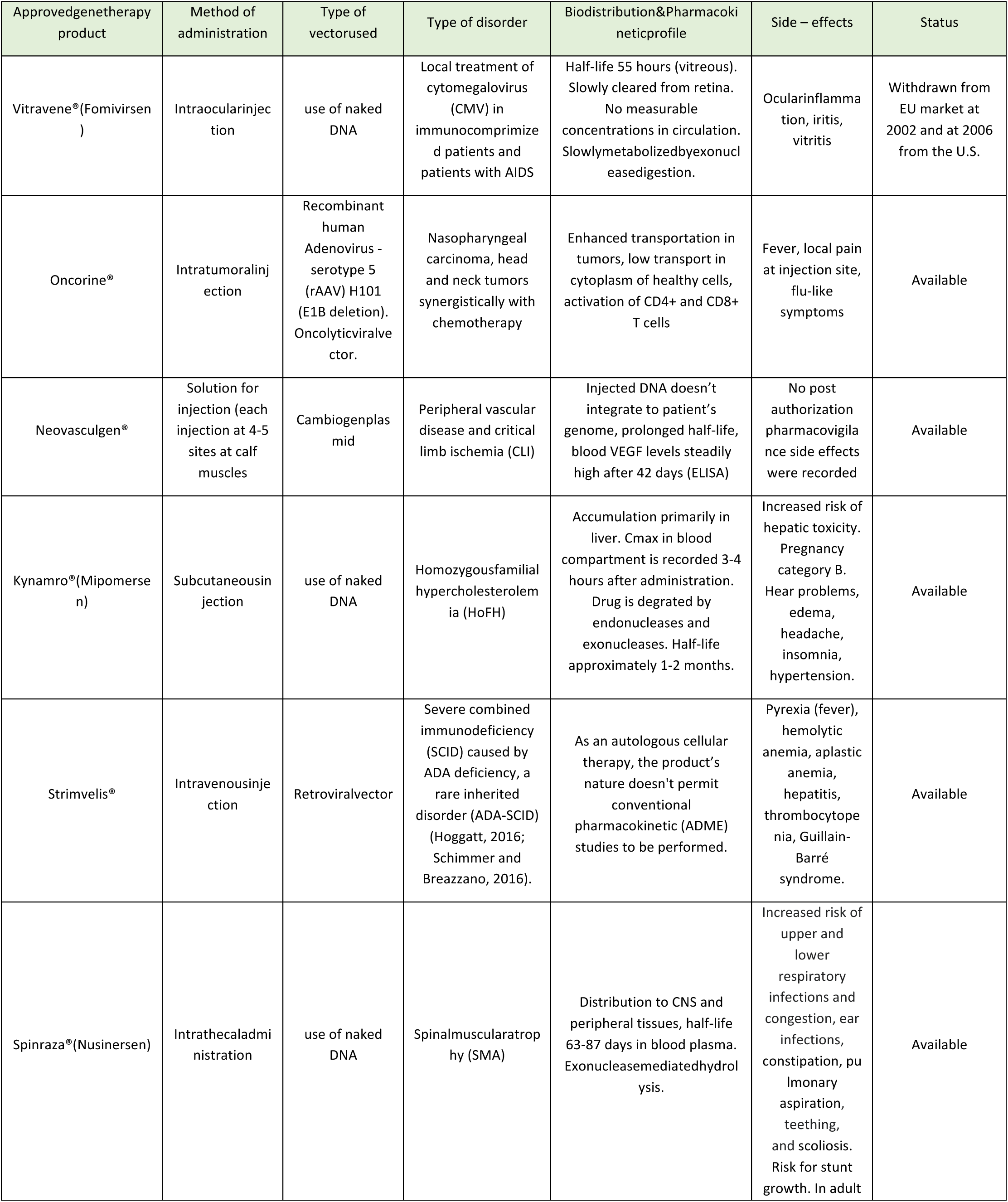
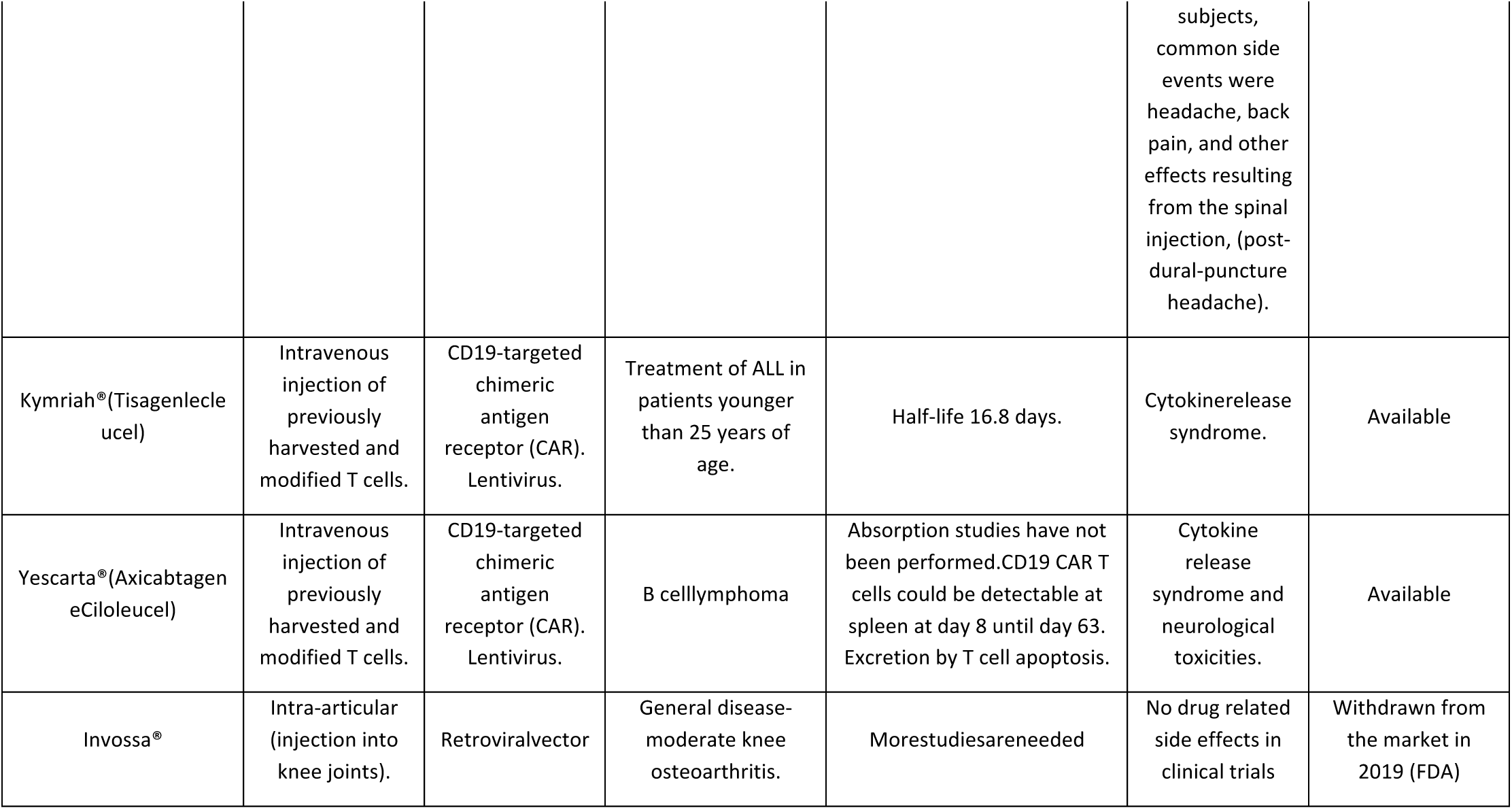
Approved gene therapy products, pharmacological actions, and regulatory information.

| Approved gene therapy product | Method of administration | Type of vector used | Type of disorder | Biodistribution & Pharmacokinetic profile | Side – effects | Status |
| --- | --- | --- | --- | --- | --- | --- |
| Vitravene® (Fomivirsen) | Intraocular injection | use of naked DNA | Local treatment of cytomegalovirus (CMV) in immunocompromised patients and patients with AIDS | Half-life 55 hours (vitreous). Slowly cleared from retina. No measurable concentrations in circulation. Slowly metabolized by exonucleases. | Ocular inflammation, iritis, vitritis | Withdrawn from EU market at 2002 and at 2006 from the U.S. |
| Oncorine® | Intratumoral injection | Recombinant human Adenovirus - serotype 5 (rAAV) H101 (E1B deletion). Oncolytic viral vector. | Nasopharyngeal carcinoma, head and neck tumors synergistically with chemotherapy | Enhanced transportation in tumors, low transport in cytoplasm of healthy cells, activation of CD4+ and CD8+ T cells | Fever, local pain at injection site, flu-like symptoms | Available |
| Neovasculgen® | Solution for injection (each injection at 4-5 sites at calf muscles) | Cambiogen plasmid | Peripheral vascular disease and critical limb ischemia (CLI) | Injected DNA doesn't integrate to patient's genome, prolonged half-life, blood VEGF levels steadily high after 42 days (ELISA) | No post authorization pharmacovigilance side effects were recorded | Available |
| Kynamro® (Mipomersen) | Subcutaneous injection | use of naked DNA | Homozygous familial hypercholesterolemia (HoFH) | Accumulation primarily in liver. Cmax in blood compartment is recorded 3-4 hours after administration. Drug is degraded by endonucleases and exonucleases. Half-life approximately 1-2 months. | Increased risk of hepatic toxicity. Pregnancy category B. Hearing problems, edema, headache, insomnia, hypertension. | Available |
| Strimvelis® | Intravenous injection | Retroviral vector | Severe combined immunodeficiency (SCID) caused by ADA deficiency, a rare inherited disorder (ADA-SCID) (Hoggatt, 2016; Schimmer and Breazzano, 2016). | As an autologous cellular therapy, the product's nature doesn't permit conventional pharmacokinetic (ADME) studies to be performed. | Pyrexia (fever), hemolytic anemia, aplastic anemia, hepatitis, thrombocytopenia, Guillain-Barré syndrome. | Available |
| Spinraza® (Nusinersen) | Intrathecal administration | use of naked DNA | Spinal muscular atrophy (SMA) | Distribution to CNS and peripheral tissues, half-life 63-87 days in blood plasma. Exonuclease mediated hydrolysis. | Increased risk of upper and lower respiratory infections and congestion, ear infections, constipation, pulmonary aspiration, teething, and scoliosis. Risk for stunt growth. In adult subjects, common side events were headache, back pain, and other effects resulting from the spinal injection, (post-dural-puncture headache). | Available |
| Kymriah®(Tisagenlecleucel) | Intravenous injection of previously harvested and modified T cells. | CD19-targeted chimeric antigen receptor (CAR). Lentivirus. | Treatment of ALL in patients younger than 25 years of age. | Half-life 16.8 days. | Cytokine release syndrome. | Available |
| Yescarta®(Axicabtagene Ciloleucel) | Intravenous injection of previously harvested and modified T cells. | CD19-targeted chimeric antigen receptor (CAR). Lentivirus. | B cell lymphoma | Absorption studies have not been performed. CD19 CAR T cells could be detectable at spleen at day 8 until day 63. Excretion by T cell apoptosis. | Cytokine release syndrome and neurological toxicities. | Available |
| Invossa® | Intra-articular (injection into knee joints). | Retroviral vector | General disease-moderate knee osteoarthritis. | More studies are needed | No drug related side effects in clinical trials | Withdrawn from the market in 2019 (FDA) |

**Table 3.**
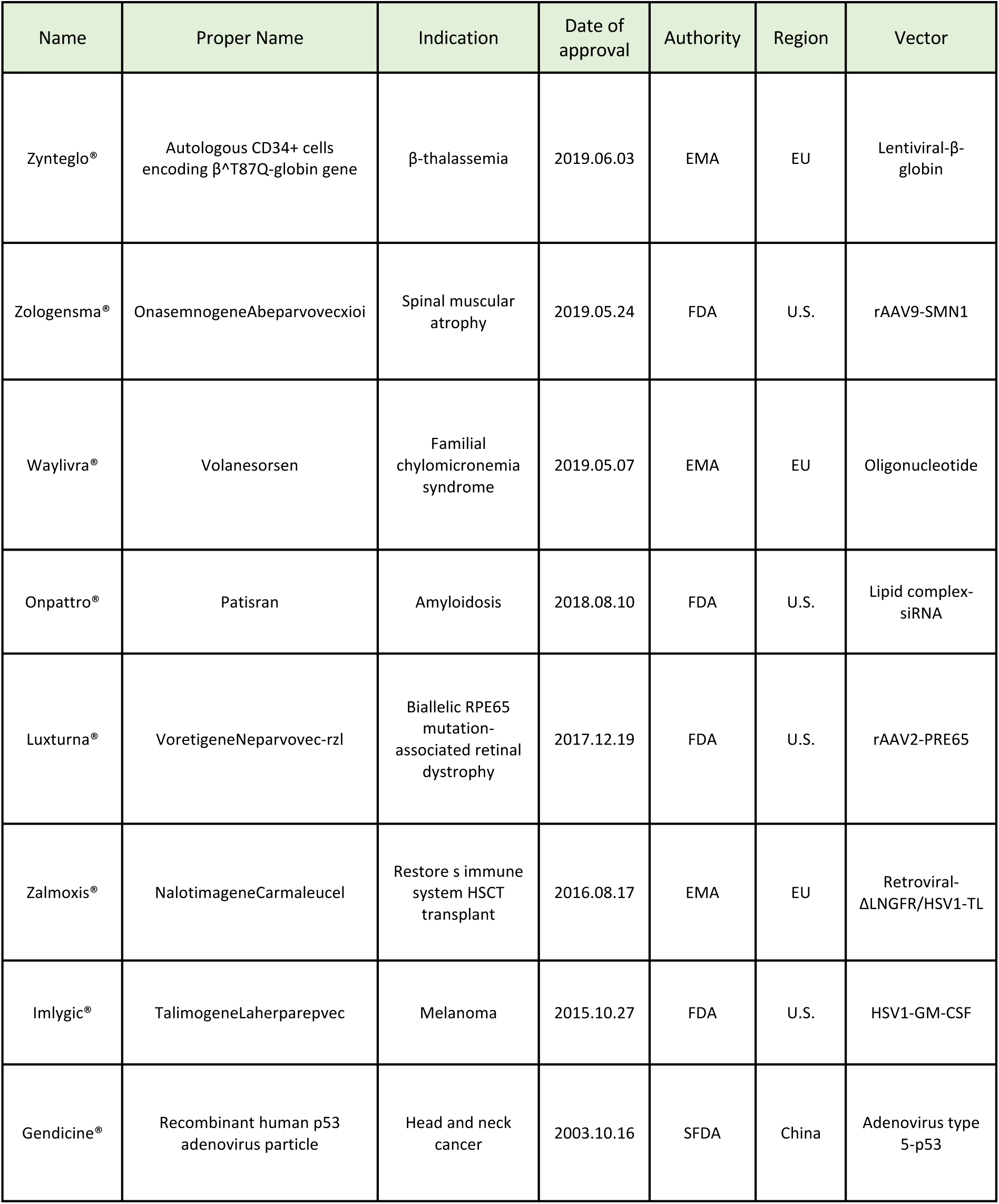
Examples of approved gene therapy products in the EU, US, and China.

In addition, the scientific community is beginning to explore other potential therapeutic indications for gene therapy. These applications include: i) Immunization against infectious diseases and cancer cells that have been mutated, ii) Synthesis of relevant protein molecules, iii) Use of RNA interference (RNAi) in gene therapy applications against cancer by inhibiting or silencing (using siRNA in silencing) genes associated with carcinogenesis and mutagenesis [22], and (iv) a cost-effective platform for the delivery of biologics (drugs derived from a biological source) [44].

Great progress has already been made in the study of diseases such as retinoblastoma and age-related macular degeneration (AMD). In both cases, therapeutic nucleic acid transfection with an adenoviral vector is being investigated, and crucial data on safety and efficacy have been obtained following the completion of phase I preclinical studies [17]. The importance of this research, as well as advances in other genetic pathologies related to retinal degradation, cannot be overstated, considering that the prevalence in the population may reach 1 in 3000 people. Several recombinant adenoviral vectors have recently been investigated in proof-of-concept studies for the treatment of many single-gene retinal disorders, although these efforts have only recently intensified following the successful outcome of the Liver Congenital Amaurosis study (focusing on the RPE65 gene) [14, 17]. Finally, diseases associated with major metabolic and skeletal disorders, as well as diseases of the central nervous system (brain and spinal cord) are being thoroughly investigated, with promising preliminary results, and recently a successful gene transfer to respiratory epithelial cells was achieved [14, 17].

Despite the above claims and the emerging trend in gene therapy, the development of novel transfectants with improved, tailored properties is crucial to further expand the scope of the technique and produce drugs with clinically meaningful outcomes. Another point to consider is that the properties and nature of the vectors we create will change greatly as they differentiate and modify, necessitating the use of several different techniques to properly characterize them. This statement is simple to understand and stems from the fact that there are no defined reference materials against which to measure the outcome of the tested material. Consequently, we need not only case-by-case evaluation and characterization of different formulations, but also the ability to control variability between different batches of the same product [4, 13]. Work on solving such problems will dramatically improve the ease of manufacturing and characterization, as well as inter-laboratory communication and duplication of data, making gene therapy products much easier to evaluate and the approval process unrivaled in speed [4, 6, 13].

## Data Availability

All data produced in the present work are contained in the manuscript

## Declarations

### Availability of data and material

Not applicable

### Code availability

Not applicable

### Ethics approval

Not applicable

